# Measuring Health System Operational Readiness for Paediatric Mass Casualty Incidents: A Scoping Review Protocol

**DOI:** 10.1101/2025.07.16.25331643

**Authors:** Jim Hickson, Wayne Thompson, Martin McNamara

**Affiliations:** School of Nursing, Midwifery and Health Systems, University College Dublin; Children’s Health Ireland at Temple Street; School of Nursing, Midwifery and Health Systems, University College Dublin

**Keywords:** emergency medicine, mass casualty incidents, operational readiness, paediatrics, disaster planning

## Abstract

**Objective:** The objective of this scoping review is to understand the frameworks and methodologies used in assessing the operational readiness (OPR) of health systems for mass casualty incidents (MCI) involving children.

**Introduction:** Children are at a higher risk of morbidity and mortality because of mass casualty incidents. A lack of operational readiness for mass casualty incidents reduces the capacity of the health system to adequately respond swiftly and safely. Understanding the methods by which health system OPR can be assessed in relation to MCIs is key to the development of recommendations for improving OPR.

**Inclusion criteria:** This review will include published evidence (including grey literature) which investigates methods of assessing health system OPR for mass casualty incidents. Studies published in the English language will be included and there will be no time limit applied to the search.

**Methods:** The search databases will consist of PubMed, Embase, CINAHL, Scopus, Global Health, ProQuest Dissertations and Theses, PsycInfo and the Cochrane CENTRAL Register of Controlled Trials. The PRISMA extension for scoping reviews will be used to allow for clarity, replicability and transparency and the review will follow the Joanna Briggs Institute (JBI) methodology for scoping reviews. Screening will be performed using Endnote 20 and Covidence software programs. Study characteristics, methodologies, assessment tools and outcomes will be extracted for analysis. A descriptive analysis, thematic synthesis, evidence mapping and a narrative synthesis will be undertaken to provide a summary of the review findings.

## Introduction

Children constitute a significant portion of Ireland’s population, representing 23.2% in 2023, the highest proportion of children in the European Union in 2022 (Department of Children, Equality, Disability, Integration and Youth, 2024). This demographic trend underscores the importance of ensuring that health systems are adequately prepared to meet the specific needs of children during emergencies, including mass casualty incidents (MCIs). Due to their unique physiological, anatomical, and psychological characteristics, children face increased risks during MCIs. Their developmental stage, dependency on adults, differing responses to trauma and medications and communication limitations make them especially vulnerable to morbidity and mortality in MCIs (Burke et al., 2010; Mason & Anderson, 2009; Chiu et al., 2022; Tovar et al., 2022).

Mass casualty incidents, defined as events that generate more patients than available resources can handle, pose a significant challenge to health systems globally (English et al, 2024). Whether as a result of natural disasters, pandemics, terrorist attacks or industrial accidents, MCIs demand swift, well-coordinated responses across multiple sectors of the health system (English et al, 2024). These events often expose underlying weaknesses in emergency preparedness, especially when it comes to paediatric care (Jenner & Piscitelli, 2024). Despite widespread recognition of children’s increased vulnerability, the health system’s readiness to respond effectively to paediatric-specific needs during MCIs remains inconsistently addressed in research (Jenner & Piscitelli, 2024).

Health system operational readiness (OPR) refers to the capacity of a health system to plan for, respond to and recover from emergencies while maintaining essential services (English et al., 2024; Kidd & Howe, 2014; Pesigan et al., 2020). It encompasses workforce readiness, resource availability, governance structures, communication protocols and coordination mechanisms (English et al., 2024; Kidd & Howe, 2014; Pesigan et al., 2020). OPR is an increasingly important concept in emergency management, especially in the context of global public health threats (English et al, 2024). However, operational readiness for MCIs involving children remains an underexplored area of research, particularly when considered through a systems-level lens. While general OPR frameworks exist, including those developed by the World Health Organization (WHO) and other international bodies, these are often adult-centric and may not adequately incorporate the distinct needs of paediatric populations (McKinnon et al, 2024). For example, surge capacity calculations, triage protocols, medical supply chains, and emergency communication strategies frequently assume adult baseline parameters, which can result in suboptimal care when applied to children (Jenner & Piscitelli, 2024). Paediatric considerations such as age-specific triage tools, child-appropriate medical supplies, psychosocial support for both children and their caregivers and staff training in paediatric care are essential yet frequently overlooked elements of MCI preparedness and response (Jenner & Piscitelli, 2024).

A preliminary search of key databases (MEDLINE, the Cochrane Database of Systematic Reviews and JBI Evidence Synthesis) revealed no current or ongoing reviews specifically addressing how health systems assess operational readiness for MCIs involving children. This gap in the literature identifies a critical need for a comprehensive synthesis of existing evidence. Although there are individual studies that address certain components of paediatric MCI response such as triage accuracy, paediatric trauma care training and disaster simulation exercises, these are often fragmented and not systematically integrated into broader discussions of health system readiness (Jenner & Piscitelli, 2024).

Scoping reviews are a valuable methodological approach when the purpose is to map existing literature on a topic, identify key concepts and highlight knowledge gaps (Munn et al, 2018). Unlike systematic reviews that focus on answering specific, narrowly defined research questions, scoping reviews are ideal for exploring complex and heterogeneous concepts such as health system preparedness (Munn et al, 2018). Given the diversity of frameworks, tools, and definitions used in the literature, a scoping review is well-suited to synthesize how paediatric OPR has been conceptualised and investigated globally (English et al, 2024).

This scoping review aims to explore the methods used to assess health system operational readiness for MCIs involving children. Specifically, the objectives are: (1) to identify and map existing frameworks used to assess OPR in paediatric MCIs, (2) to understand how health systems have evaluated their readiness for such events and (3) to highlight gaps and limitations in the existing research base. By doing so, the review will not only inform future research but also support policy development and planning in health systems where paediatric preparedness is essential. Understanding the operational readiness of health systems for paediatric MCIs is critical for several reasons. Firstly, children have different clinical and psychosocial needs during emergencies, which require specialised training, resources and protocols (Jenner & Piscitelli, 2024). Secondly, health systems that lack paediatric-specific emergency planning may experience increased mortality and morbidity among children during disasters (Jenner & Piscitelli, 2024). Thirdly, the aftermath of an MCI often includes long-term consequences such as trauma, displacement and chronic health conditions that disproportionately affect younger populations (Uddin et al, 2024).

Despite the recognition of these factors, several limitations persist in the current literature. Many studies focus on singular aspects of preparedness such as triage accuracy or equipment availability rather than providing a holistic systems-level perspective (Mourid et al, 2025). Furthermore, there is inconsistency in how OPR is defined and measured across studies and regions, making comparisons difficult (English et al, 2024). Few studies employ validated assessment tools or frameworks tailored to paediatric needs. Grey literature, including government reports and internal health system assessments, may contain valuable insights that are not captured in academic publications, highlighting another potential gap. There is also a geographic and contextual imbalance in the literature. Much of the available research originates from high-income countries, particularly the United States, which may limit the applicability of findings to other settings, including Ireland (Burke et al, 2010). This scoping review seeks to address these limitations by employing a comprehensive search strategy that includes peer-reviewed and grey literature, with a focus on identifying diverse perspectives and contextual factors. The review will be guided by established methodological frameworks, including the JBI methodology for scoping reviews and the PRISMA-ScR checklist, ensuring transparency, rigour, and reproducibility.

Ultimately, the findings from this review will have significant implications for both research and policy. By identifying what is currently known and where the gaps lie, the review will support the development of more robust, child-centred preparedness frameworks and encourage the integration of paediatric considerations into health system planning. Policymakers, emergency planners, clinicians and researchers will benefit from a clearer understanding of how to strengthen operational readiness for MCIs involving children.

Review question

Using the PCC framework for research questions, the question for this scoping review is: “what methods have been used to assess health system (P) operational readiness (C) for mass casualty incidents involving children (C)?”. The review will aim to address questions:

- How has the capacity of health systems to respond to paediatric MCIs been assessed?
- What frameworks or methodologies have been used to assess OPR in this context?
- What gaps exist in OPR for paediatric MCIs and what improvements are needed?

## Inclusion criteria

### Participants

Studies will be included where the participants consist of health systems or healthcare institutions. Studies focused on non-human health systems, e.g. wildlife impact of MCIs/disasters, will be excluded from the review.

### Concept

For this scoping review, operational readiness is defined (English et al, 2024) as the capacity of a health system to effectively prepare for, respond to and recover from the relevant incident. This will include planning and preparedness, resource allocation, surge capacity, coordination, communication and response mechanisms. Operational readiness emphasises a proactive approach to ensure an adequate response to the crisis. Studies will be included where the focus is assessing any of these components.

### Context

The context of studies for inclusion in this review will be mass casualty incidents There will be no exclusion based on geographical location of studies. Studies where the context is solely infectious disease/pandemics without a discussion of mass casualty readiness will be excluded. Studies will not be limited to those with a focus on paediatric MCIs. It was decided that by limiting to those focusing on a paediatric population, potentially valuable data may be missed.

### Types of sources

This scoping review will include all study designs, including randomised and non-randomised controlled trials, observational studies, cohort studies, case-control studies and analytical cross-sectional studies. Additionally, descriptive observational study designs and systematic reviews will be considered for inclusion. Grey literature, including guidelines, protocols, legislation and frameworks sourced via OpenGrey and ProQuest, will be considered for inclusion. Both national and international grey literature will be included, acknowledging the lack of evidence available specifically focusing on the Irish context.

## Methods

The proposed scoping review will be conducted in accordance with the JBI methodology for scoping reviews (Peters et al, 2020).

### Search strategy

This search strategy will aim to source all available evidence. An initial basic search was completed of CINAHL and Embase databases. The key words and phrases used in the abstracts of the sourced articles and the index terms were then used to formulate a search strategy for PubMed, Embase, Medline, CINAHL, Scopus, Global Health, PsycInfo and ProQuest Dissertations and Theses (See appendix 1). The search strategy was then adapted for each relevant database. A backward citation search will be completed to identify any other relevant publications that were not already identified in the search. Studies published in the English language will be included. Studies in other languages will be excluded. However, sentinel studies identified during reverse citation searches may be translated using Google Translate. Studies published between 01/01/2015 and 01/05/2025 will be included. This date restriction was chosen for feasibility of the review and it was felt that sentinel papers published prior to this date range will be identified through reverse citation searching..

## Data Availability

All data produced in the present study are available upon reasonable request to the authors

## Study/Source of evidence selection

All records retrieved from the search strategy will be imported to EndNote20 for data management and secure record keeping. Records will then be uploaded to Covidence software, where duplicates will be removed. Titles and abstracts will be screened by two independent reviewers to assess for their inclusion based on the inclusion and exclusion criteria listed above. Sources that are considered potentially eligible will be forwarded for full text review. Where there are disagreements between reviewers regarding the eligibility of a record, a third independent reviewer will make the final decision regarding their eligibility. Any reasons for excluding articles will be documented for reporting in the review. The results of this search will be reported in full in the final review. They will be displayed using a PRISMA flow diagram (PRISMA, 2018).

### Data extraction

Following the screening process, the resultant articles will be included for data extraction using an author-developed data extraction tool (Appendix II). This data extraction tool will be piloted on 10 articles from the initial search and iteratively refined to ensure all relevant data will be included. The data extracted will include study characteristics, methodologies, frameworks, study context and outcomes/key findings which pertain to the review question. If any data is missing or requiring clarification, the study authors will be contacted where appropriate. If data is still missing this will be noted in the review to maintain transparency.

### Data analysis and presentation

A descriptive analysis will be completed which will provide a synthesis of the characteristics of the included articles. A thematic synthesis will be performed to identify any overarching themes, frameworks and key findings related to the review question. Evidence mapping will allow the identification of any evidence gaps and provide recommendations for further research. The results will be presented in a tabular format

## Acknowledgements

This review is to contribute towards the PhD award for author JH.

## Declarations

The authors have no declarations.

## Author contributions

All authors contributed equally to the development of the search strategy, the screening and data extraction processes. JH performed the analysis and composition of the manuscript with iterative feedback from the remaining authors.

## Ethics Statement

As this review will use data from publicly available sources, ethical approval is not required.

## Appendices

### Appendix I: Search strategy

#### 1. PubMed

“disaster*” OR “mass casualt*” OR “terrorism” OR “terrorist attack*” OR “mass shooting*” OR “accident*” OR “bioterror*” OR “emergencies”[MeSH] OR “disasters”[MeSH] OR “mass casualty incidents”[MeSH] OR “terrorism”[MeSH] OR “bombs”[MeSH]

AND

“operational readiness” OR “preparedness” OR “surge planning” OR “surge capacity” OR “disaster planning” OR “disaster preparedness” OR “disaster relief planning” OR “disaster response” OR “Mass casualty response*” OR “performance” OR “response capacity” OR “disaster planning”[MeSH]

Limited to title and abstract, published in the English language, published in the last 10 years. Test search on 07/03/2025 resulted in 24,409 articles.

#### 2. Embase

(‘emergency’:ab,ti OR disaster:ab,ti OR ‘mass casualty’:ab,ti OR ‘mass casualty incident’:ab,ti OR ‘mass casualties’:ab,ti OR ‘terrorism’:ab,ti OR ‘terrorist attack’:ab,ti OR ‘terrorist attacks’:ab,ti OR ‘mass shooting’:ab,ti OR ‘mass shootings’:ab,ti OR ‘accident’:ab,ti OR ‘bioterror’:ab,ti OR ‘bioterrorism’:ab,ti) AND (‘operational readiness’:ab,ti OR ‘preparedness’:ab,ti OR ‘surge planning’:ab,ti OR ‘surge capacity’:ab,ti OR ‘disaster planning’:ab,ti OR ‘disaster preparedness’:ab,ti OR ‘disaster relief planning’:ab,ti OR ‘disaster response’:ab,ti OR ‘mass casualty response’:ab,ti OR ‘response capacity’:ab,ti)

Limited to title and abstract, published in the English language, published in the last 10 years.

*Test search 07/03/2025: 7,640 results*

#### 3. CINAHL

(“emergencies” OR “disasters” OR “disaster” OR or “mass casualty” OR “mass casualty incident” OR “mass casualties” OR “terrorism” OR “terrorism” OR”terrorist attack” OR “terrorist attacks” OR “mass shooting” OR “mass shootings” OR “accident” OR “bioterror”)

AND

(“operational readiness” OR “preparedness” OR “surge planning” OR “surge capacity” OR “disaster planning” OR “disaster planning” OR “disaster preparedness” OR “disaster relief planning” OR “disaster response” OR “Mass casualty response” OR “response capacity”)

Limited to title and abstract, published in the English language, published in the last 10 years.

*Test search 07/03/2025: 3,339 results.*

#### 4. Scopus

TITLE-ABS-KEY({emergencies} OR {disasters} OR {disaster} OR {mass casualty} OR {mass casualty incident} OR {mass casualties} OR {terrorism} OR {terrorist attack} OR {terrorist attacks} OR {mass shooting} OR {mass shootings} OR {accident} OR {bioterror})

AND

TITLE-ABS-KEY({operational readiness} OR {preparedness} OR {surge planning} OR {surge capacity} OR {disaster planning} OR {disaster planning} OR {disaster preparedness} OR {disaster relief planning} OR

{disaster response} OR {Mass casualty response} OR {response capacity}) Limited to title, abstract and key-words, published in the English language.

*Test search 07/03/2025: 22,521 results*

#### 5. Global Health

(“emergencies” OR “disasters” OR “disaster” OR or “mass casualty” OR “mass casualty incident” OR “mass casualties” OR “terrorism” OR”terrorist attack” OR “terrorist attacks” OR “mass shooting” OR “mass shootings” OR “accident” OR “bioterror”)

AND

(“operational readiness” OR “preparedness” OR “surge planning” OR “surge capacity” OR “disaster planning” OR “disaster planning” OR “disaster preparedness” OR “disaster relief planning” OR “disaster response” OR “Mass casualty response” OR “response capacity”)

Limited to abstract, published in the English language, published in the last 10 years. Test search 07/03/2025: 7 results*

#### 6. ProQuest Dissertations and Theses

(“emergencies” OR “disasters” OR “disaster” OR or “mass casualty” OR “mass casualty incident” OR “mass casualties” OR “terrorism” OR “terrorism” OR”terrorist attack” OR “terrorist attacks” OR “mass shooting” OR “mass shootings” OR “accident” OR “bioterror”)

AND

(“operational readiness” OR “preparedness” OR “surge planning” OR “surge capacity” OR “disaster planning” OR “disaster planning” OR “disaster preparedness” OR “disaster relief planning” OR “disaster response” OR “Mass casualty response” OR “response capacity”)

Limited to abstract and summary text, published in English language, published in last 10 years.

*Test search 07/03/2025: 1,025 results*

#### 7. Cochrane CENTRAL Register of Controlled Trials

(“emergencies” OR “disasters” OR “disaster” OR “mass casualty” OR “mass casualty incident” OR “mass casualties” OR “terrorism” OR “terrorist attack” OR “terrorist attacks” OR “mass shooting” OR “mass shootings” OR “accident” OR “bioterror”)

AND

(“operational readiness” OR “preparedness” OR “surge planning” OR “surge capacity” OR “disaster planning” OR “disaster planning” OR “disaster preparedness” OR “disaster relief planning” OR “disaster response” OR “Mass casualty response” OR “response capacity”)

*Test search 07/03/2025: 8 results*

#### 8. PsycInfo

(“emergencies” OR “disasters” OR “disaster” OR “mass casualty” OR “mass casualty incident” OR “mass casualties” OR “terrorism” OR “terrorism” OR “terrorist attack” OR “terrorist attacks” OR “mass shooting” OR “mass shootings” OR “accident” OR “bioterror”)

AND

(“operational readiness” OR “preparedness” OR “surge planning” OR “surge capacity” OR “disaster planning” OR “disaster planning” OR “disaster preparedness” OR “disaster relief planning” OR “disaster response” OR “Mass casualty response” OR “response capacity”)

Limited to abstract, published in the English language, published in the last 10 years.

*Test search 07/03/2025: 1,040 results*

### Appendix II: Data extraction tool template

|  |
| --- |
| DOI |
| Title |
| Year |
| Country/region |
| Aim |
| Study design |
| Population |
| Inclusion criteria |
| Exclusion criteria |
| Total number of participants |
| Type of MCI |
| Scale<br>(regional/national/international) |
| Tools/framework used |
| Summary |
| Recommendations |
| Gaps identified |
| Paediatric specific findings |
| Limitations |
| Notes |
| Themes |

